# A HEURISTIC MODEL FOR SPREAD OF COVID-19 INFECTION CASES IN INDIA

**DOI:** 10.1101/2020.04.25.20079483

**Authors:** Bishwajit Bhattacharjee

**Affiliations:** Emiretus Professor, Department of civil engineering, Indian Institute of Technology Delhi, Delhi, India-110016.

**Keywords:** Heuristic model, COVID-19, Spread of infection, Indian cases

## Abstract

A simple heuristic model for spread of COVID-19 infections in India is presented and compared with reported data up to April 10, 2020. Spread of infection is considered to be initiating from infected individuals and spread is assumed to take place in a compounding manner. Some of the data needed are taken from readily available sources in the web. The possible progress is then estimated based on model presented and possible scenarios are highlighted.

## Introduction

SIR(Susceptible, Infected, Recovered) model is commonly used to predict growth of epidemic does not seem to be readily applicable in the case of spread od COVID-19 infections in India. This is because the basic assumption made in the model that total population under consideration does not change with time is not valid under the current Indian circumstances. Consequently, the rate of change total population, comprising of susceptible population S(t), infected I(t) and recovered (or dead) population R(t) at any time t, with time is not zero. Mathematically,

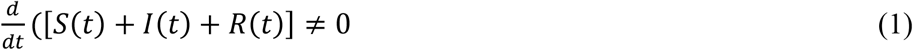

This is because of situation in India, with large population dispersed all over the country, having several nucleation points, e.g., Jaipur, Kerala, Maharashtra, Punjab, Delhi etc. These points are scattered all over the country, hence new population get included in susceptible set and thus total population varies with time. The SIR model’s other assumption of interaction i.e. ds/dt=-bS(t)I(t) with negative sign implies that S(t) reduces with time is also not valid. In India, new nucleation points are created by import of infected person, at times unidentified, from inside and outside the country and freely interacting with others till discovered. On the other hand a simple model conceptualized based on an analogy to compounding the number seems to work better. The model is described below.

## Model and estimation

If the rate of infection is R_0_, i.e. number of person on an average get infected from 1 infected person, then the number of persons infected after the incubation period would be 1+R_0_, without any external intervention. Since all these persons would cause infection to others they have interacted with, after another cycle of incubation period, the number of infected persons would be (1+R_0_)^2^ and so on, after “m” such cycles number of infected person is (1+R_0_)^m^. There is statistical uncertainty over incubation period. However, one can work with mean or expected incubation period “mIP” (time period). Thus expected number of cycle per day is 1/mIP. Hence expected number of infected person after one day is (1+R_0_)^(1/mIP).^ After n days the expected number of infected cases is (1+R_0_)^(n/mIP).^ Spread of the infection occurred in different locations as nucleation points, also the initiation of spread would have occurred at different time, hence a more general equation for number of infected cases I(n) without external intervention, would be given by

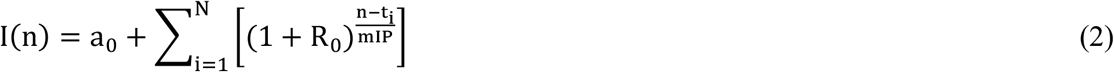

Where, n is the day, a_0_ is number of infected cases at n=0, t_i_=0 for t=1, before actual spread started and N is the number of nucleation locations with one initial infected person. The mIP can be assumed from reported statistical studies. a_0_ in this case is 3 as this is the number of infected people throughout the initial days of reporting during the month of February 2020. Commonly the accepted range of incubation period starts from 2 days. Some study report that 97.5% of the time the number of days is lower than 11.5 days and the median at 5.1 days, i.e. 50% of the time the incubation period is less than 5.1 days[1-3]. Others tend to report the mean incubation period close to 5 days. However, it is clear that the distribution is skewed towards the left and possibly the distribution fits best to a 2 parameter β distribution. Values of α and β of the distribution are likely to be close to 2 and 6 respectively and the mean is likely to be slightly higher than 5.1 days. In absence of more information mIP is assumed as 5.5 days. Estimation of t_i_ can be done from the data by tracing down the sudden surge in the cases and locating actual nucleation points. However in absence of that, one can assume addition of one person as nucleation point everyday on an average basis for the whole country from the time of detected spread. With these assumptions the equation (2) can be further simplified as described below. The summation terms in equation (2) can be expanded for t_i_=0, 1,2…‥N and simplified using the sum of geometrical progression series:

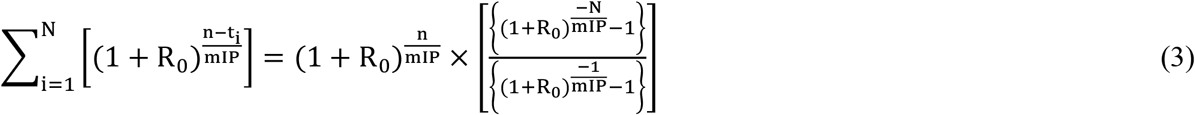

Consequently equation (2) is rewritten as equation (4) below

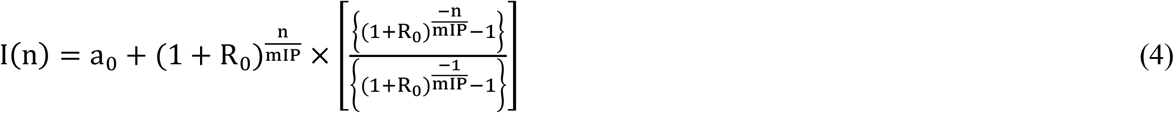

Thus I(n) on nth day from initiation can be calculated. In this case 1st March is assumed as n=0, the day of spread with a surge from initial state. However, a person as a source for spread of infection would act as source up to a maximum number of days of active source period, as after that the person would be either quarantined or hospitalized or ceased to be an active source case, and already contributed to toal number of cases. This period is assumed to be 14 days and accordingly equation (4) is modified as:

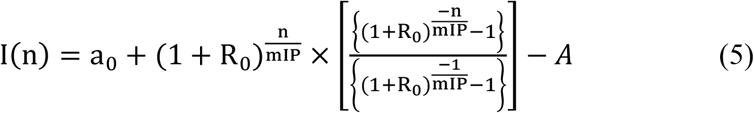

where, 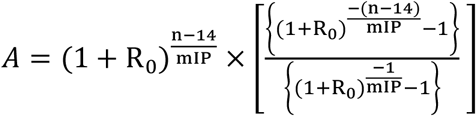 for n>14 else, A=0;

The value is R_0_ ranges from 1.5 to 2, or even to a higher value of 3. The estimated I(n) values considering R_0_ as 1.75, against actual reported in public domain [4] is plotted in Fig.1. There seems to be a broad agreement between the actual and estimated, till 36 days. So far data related to 55 days are available. The above equation 5 is obtained assuming no intervention. With intervention the rate would be lower. The intervention in the form of lock down and social distancing started from March 23, 2020, i.e n=22. Effect of lintervention seems to lag by about 14 days. Mathematically effect of intervention is considered in the next section.

**Fig 1.**
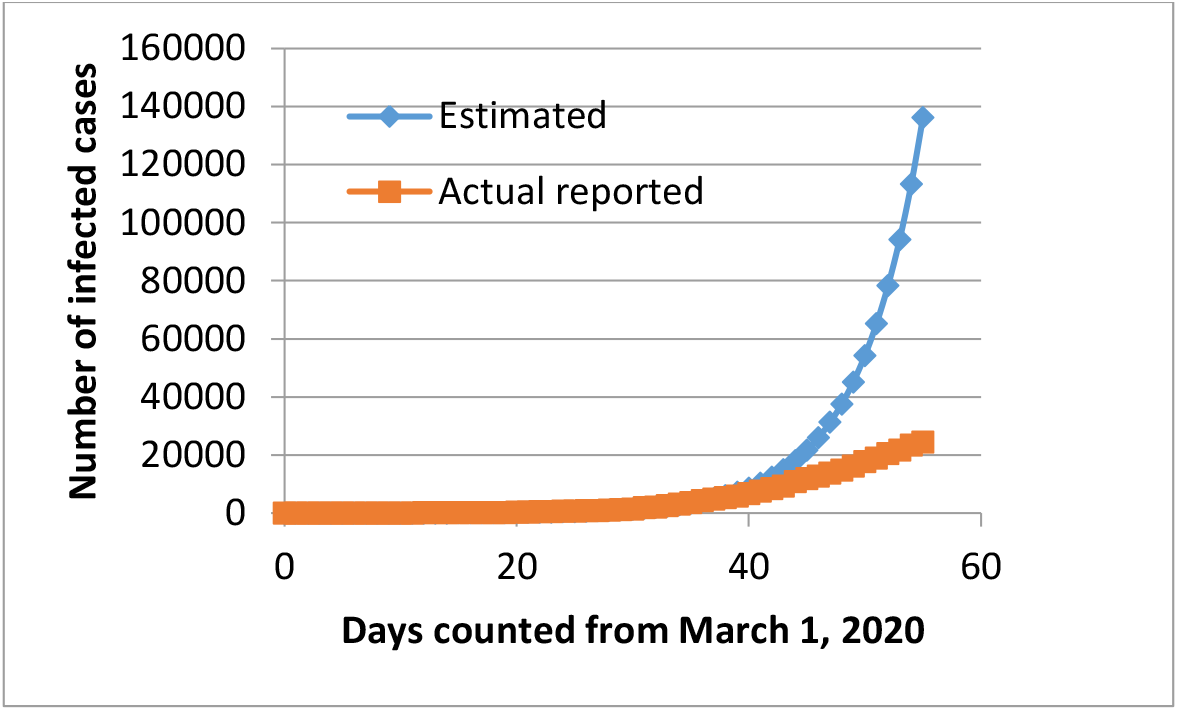
Model estimated infected cases and actual reported against days with constant initial rate

### The Effect of Lock down and social distancing Intervention

The lock down and social intervention would reduce the probability of contact of the infected person with rest of the community and hence would reduce the rate. The rate R_0_ is likely to reduce assymtotically to zero over a very long i.e. infinite period of time. The effectiveness of lock down and social distancing would be reliazed with a time lag as mentioned earlier thus R_0_ can be assumed to decrease exponentially and the new rate is R(n) is a function of day as given in the equation below.

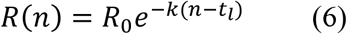

The k a constant representing effectiveness of intervention and t_l_ is the day from which effectiveness of intervention is realized. In this case t_l_=36. Coffeciant of (intervention) effectiveness would vary from society to society and so is t_l_. Thus the new equation for n≥37 is given as:

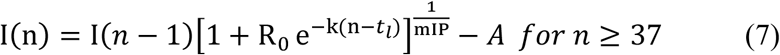

A value of k=0.081 provides a good agrrement between model and actual I(n) values as shown in Fig.2. For a a steady I(n) value In=I(n-1), putting this condition yield

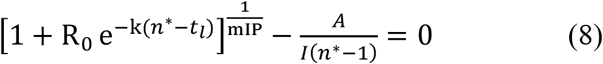

**Fig 2.**
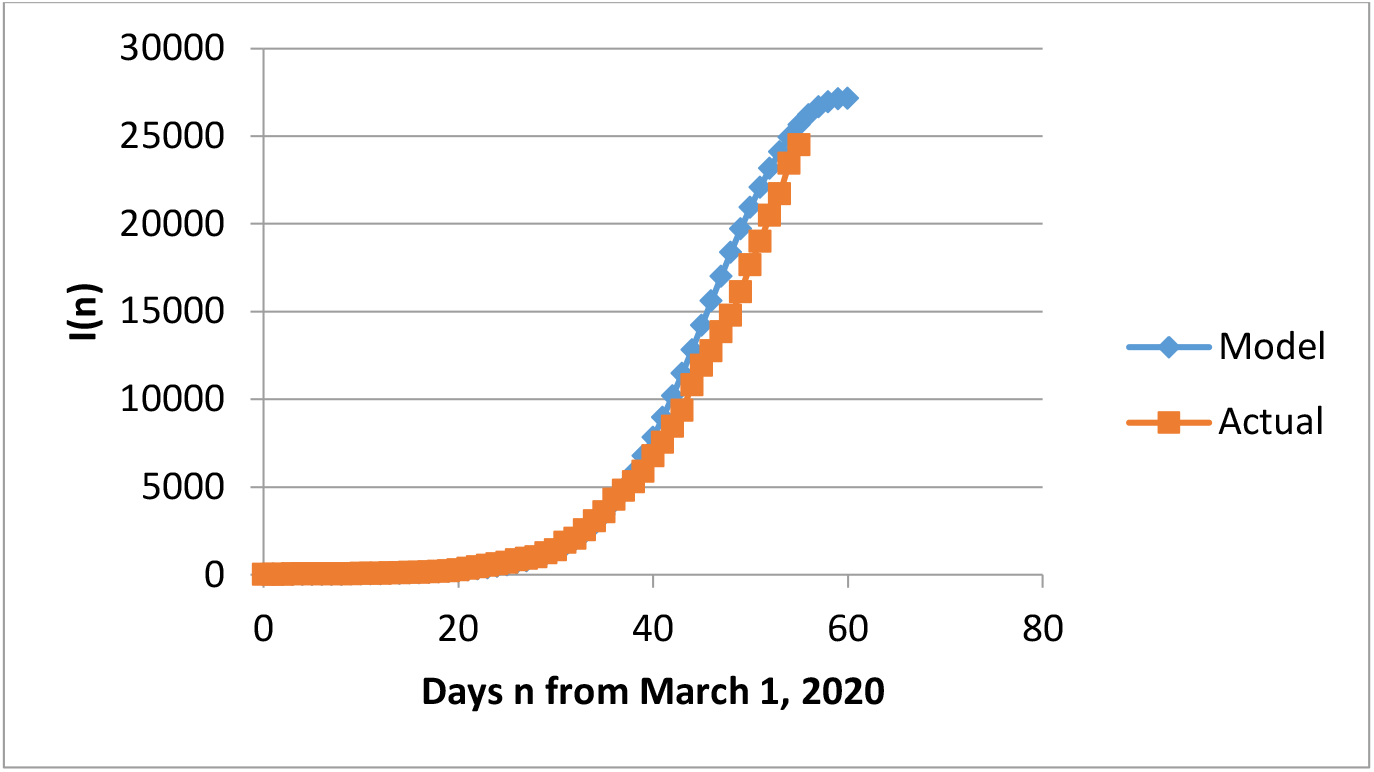
Model estimated infected cases and actual reported against days with Intervention

Solving this transcendental equation one would get n^*^≈67; which means that the growth of Covid -19 infection spread shall stabilize around 12 in India. Howevr, there are many assumption particularly of numbers in this exercise, hence one can expect that the situation may stabilize within 10-15 days. One can carryout sensitivity analysis with respect uncertain parameters and obtainpossible outcome situations.

## Concluding Remarks

The model however demonstrates that the intervention in the form of lock down and social distancing can work in reducing the rate of spread of infection. The uncertainties in the model can be reduced with more reliable data and necessary refinements

## Data Availability

DATA IS AVAILABLE FREELY

https://en.wikipedia.org/wiki/2020_coronavirus_pandemic_in_India

## REFERENCES

[1] Stephen A. Lauer, Kyra H. Grantz, Qifang Bi, Forrest K. Jones, Qulu Zheng, Hannah R. Meredith, Andrew S. Azman, Nicholas G. Reich, Justin Lessler. (2020). “The Incubation Period of Coronavirus Disease 2019 (COVID-19) From Publicly Reported Confirmed Cases: Estimation and Application”. Annals of Internal Medicine, DOI: 10.7326/M20-0504

[2] https://www.who.int/news-room/q-a-detail/q-a-coronaviruses

[3] https://www.worldometers.info/coronavirus/coronavirus-incubation-period/

[4] https://en.wikipedia.org/wiki/2020_coronavirus_pandemic_in_India

